# Latent chronic stress: identification and characterization in a preventive medicine cohort

**DOI:** 10.1101/2025.11.21.25340737

**Authors:** Maryne Lepoittevin, Marie Bringer, Maddalena Balia, Pierre Bauvin, Alaedine Benani, Franck Zenasni, Sylvain Bodard

## Abstract

Chronic stress involves both subjective appraisal and multisystem biological activation, yet these dimensions do not always converge. In a preventive medicine cohort of 1,383 adults, we investigated dissociations between perceived stress and allostatic load (AL) to identify stress phenotypes, with a particular focus on a latent profile characterized by high AL despite low perceived stress. Participants completed a comprehensive preventive health assessment and a 34-item perceived stress questionnaire (ZPSS), and AL was computed across five physiological domains. Using predefined thresholds, we delineated four stress profiles: homeostatic (low AL/low ZPSS), prodromal (low AL/high ZPSS), decompensated (high AL/high ZPSS), and a latent profile (high AL/low ZPSS). Nearly half the cohort exhibited elevated biological or psychological stress, and 14% belonged to the latent profile. These individuals showed significant multisystem dysregulation despite low perceived stress and reported the highest emotional expressivity, suggesting preserved emotional functioning despite physiological strain. Several mechanisms may underlie this dissociation, including expressive buffering, partial psychological habituation, and residual “biological scars’’ from past stress exposures. Together, these findings reveal a substantial burden of silent physiological stress in ostensibly healthy adults and highlight the need for longitudinal and multimodal approaches to identify pre-symptomatic states and guide early, emotion-informed preventive interventions.

## Introduction

Stress is an adaptive physiological response to a situation perceived as threatening or constraining. More broadly, it can be defined as a natural reaction of the body and mind to physical or psychological demands [1]. Whilst stress response is beneficial in the short term, it becomes maladaptive when it persists over time. This state, known as chronic stress, is now recognized as a major risk factor in many chronic diseases. It notably contributes to the development or aggravation of pathologies such as cardiovascular diseases [2], specific cancers [3] or Alzheimer’s disease [4]. Stress can be assessed according to several dimensions [5]. On the physiological level, the allostatic load (AL) constitutes an integrative measure of the biological impact of chronic stress. It reflects cumulative wear and tear from repeated activation of the body’s regulatory systems, such as the neuroendocrine, immune, and cardiovascular systems. The original AL included 10 biomarkers: cortisol, norepinephrine, epinephrine, dehydroepiandrosterone (DHEA), systolic blood pressure (SBP), diastolic blood pressure (DBP), waist-to-hip ratio (WHR), high-density lipoprotein (HDL), total cholesterol (TC), and glycosylated hemoglobin (HbA1C) [6, 7]. Body mass index (BMI) and C Reactive Protein (CRP) have been extensively used as well since then, as well as a number of other biomarkers [8]. Recent approaches emphasize multi-system AL scores that combine multiple physiological domains, in line with the framework proposed by McEwen and colleagues, and later refined by Palix et al, [9, 10]. In this perspective, we computed AL across five domains (hypothalamic–pituitary–adrenal axis, sympathetic/autonomic, cardiovascular, lipid–metabolic, and glyco-inflammation–renal), highlighting AL as a global, non-specific yet sensitive index of cumulative stress burden.

Conversely, perceived stress is a psychological, self-reported dimension based on an individual’s appraisal of their own stress level [11]. This appraisal is influenced by coping strategies, beliefs, and prior experiences [12]. In contrast, physiological/biological indicators of stress, such as AL and related physiological stress indices, quantify the multisystemic biological burden of stress with relative independence from personal awareness. These physiological/biological measures capture the “wear and tear” on regulatory systems, whereas self-reported (perceived) measures reflect the individual’s conscious appraisal of stress. In line with the unified framework proposed by Epel and colleagues, stress assessment benefits from integrating subjective experience with biological and behavioral readouts across time (exposure, reactivity, recovery, restoration), rather than privileging a single dimension [13]. Discrepancies between physiological/biological indicators and self-reported perceived stress suggest the existence of distinct stress profiles. Building on this framework, we define four joint AL–perceived stress profiles: a homeostatic profile (low AL, low perceived stress), a prodromal profile (low AL, high perceived stress), a latent/asymptomatic profile (high AL, low perceived stress), and a decompensated profile (high AL, high perceived stress). The detection of these latent or asymptomatic forms of chronic stress and their timely clinical management is a major public health issue. Indeed, if this stress is not identified and managed, it can promote an insidious progression of chronic pathologies, escaping classical screening approaches. Early intervention, based on lifestyle adjustments, support for emotion regulation, and targeted behavioral support, could favorably reorient the health trajectory of these individuals before the onset of organic pathologies [14].

Here, we aimed to identify and characterize this asymptomatic chronic stress phenotype by jointly assessing AL, across five biological domains using an aggregated score and perceived stress. The present study was conducted in a cohort of adults attending a dedicated preventive medicine center for comprehensive health check-ups. Our working hypothesis is that the dissociation between biological stress burden and self-reported stress, described in prior work [7, 15–18], would replicate in an independent preventive-medicine cohort. Accordingly, we sought to (i) delineate distinct stress profiles based on the joint distribution of AL and perceived stress, with a particular focus on the latent/asymptomatic subgroup, and (ii) describe their demographic and biological correlates, thereby extending previous findings by providing domain-level biological characterization and quantifying the prevalence of each profile.

## Materials and methods

### Patients

Participants were recruited within the Zoī preventive medicine cohort [19], a longitudinal study designed to investigate early biological and behavioral determinants of health in adults undergoing comprehensive preventive assessments. Recruitment occurred between November 2023 and June 2025, and all individuals who completed a full preventive assessment during this period were included in the present analysis. A total of 1,383 individuals aged ≥18 years were analyzed. No additional inclusion criteria were applied; however, participants with missing key data (perceived stress or biological measures) were excluded. Baseline characteristics of the study population are summarized in **Table 1**. The study protocol was approved by the *Comité d’Éthique pour la Recherche Biomédicale de Rabat* (CERB 159-25).

**Table 1:**
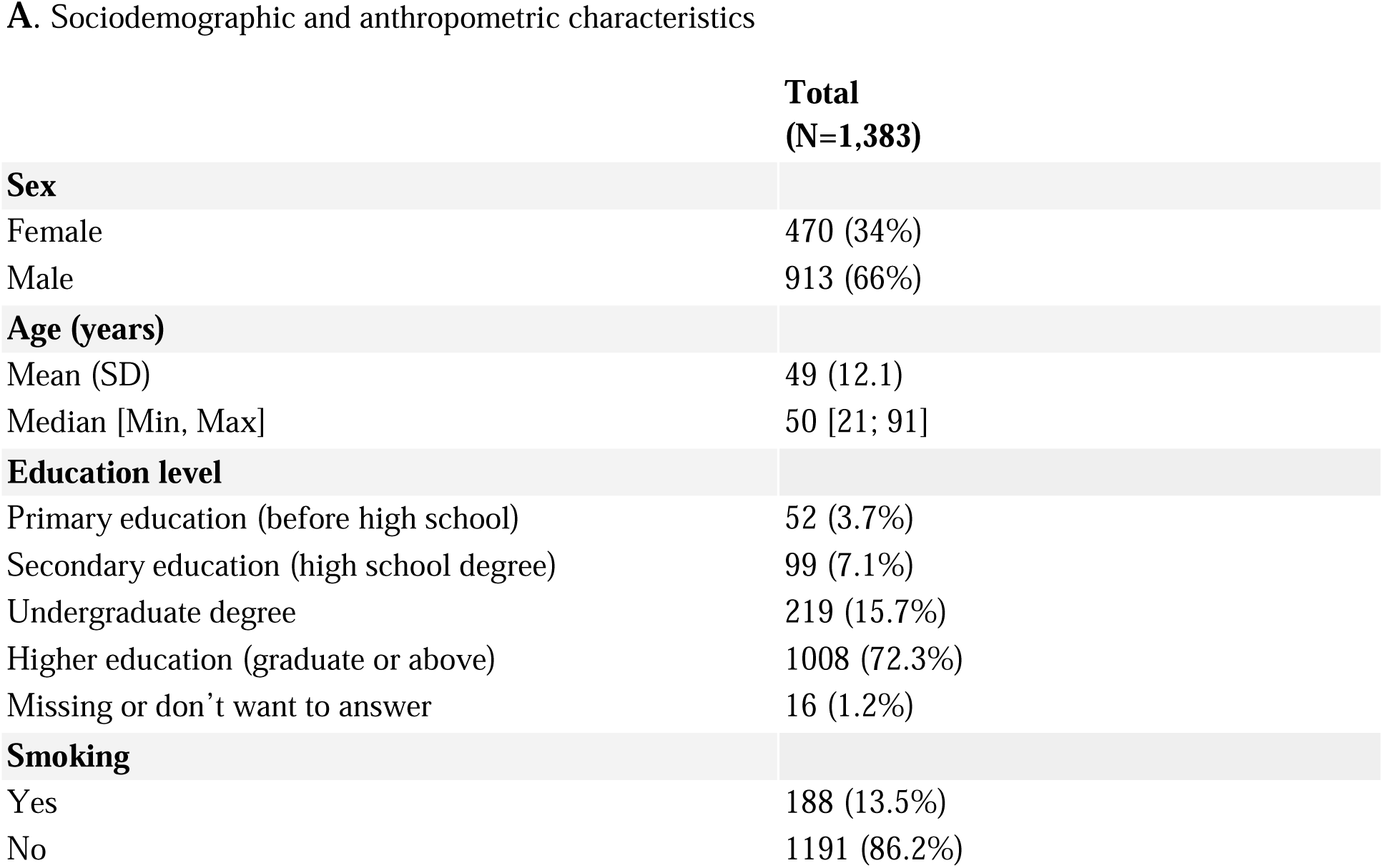
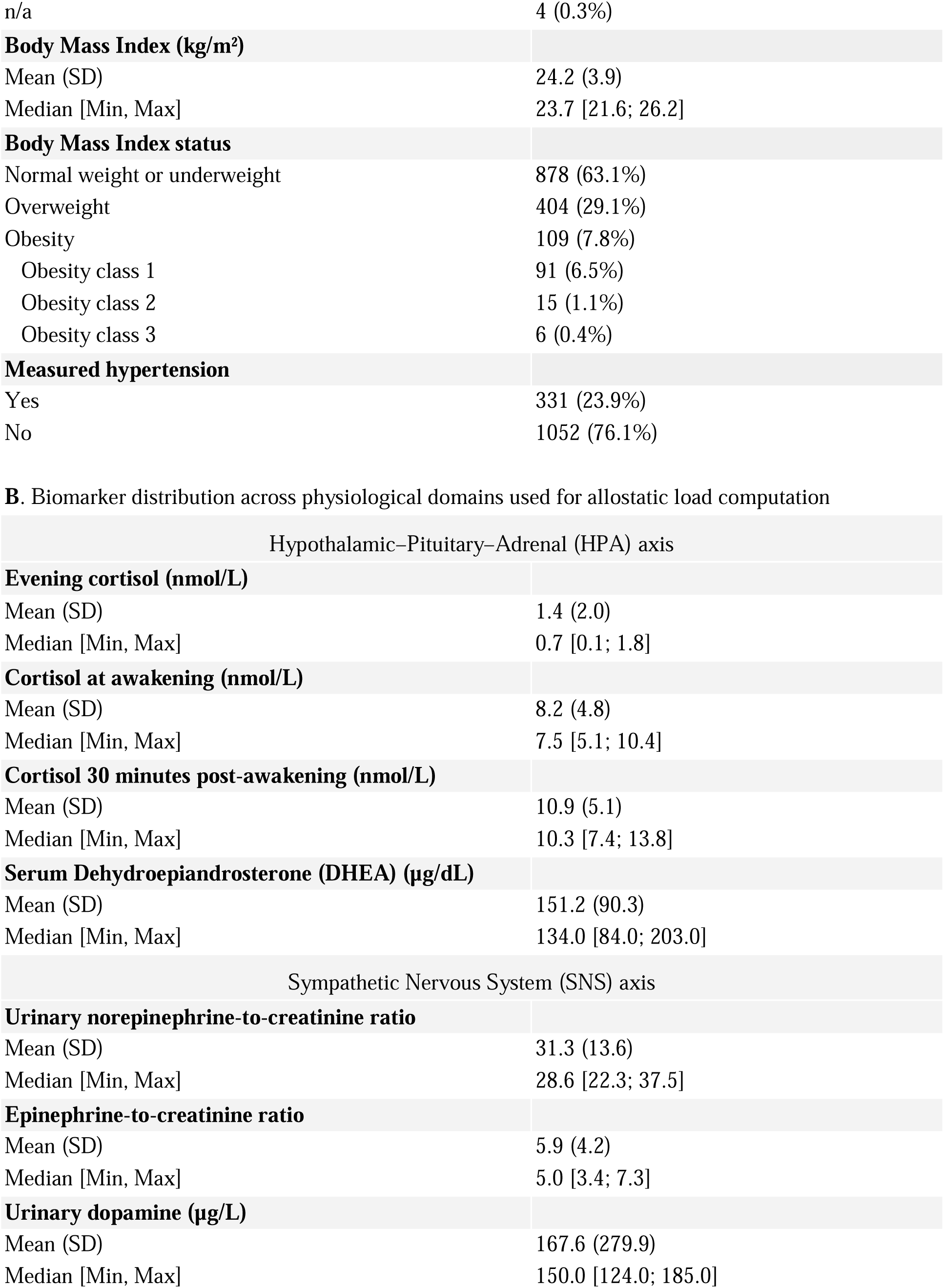
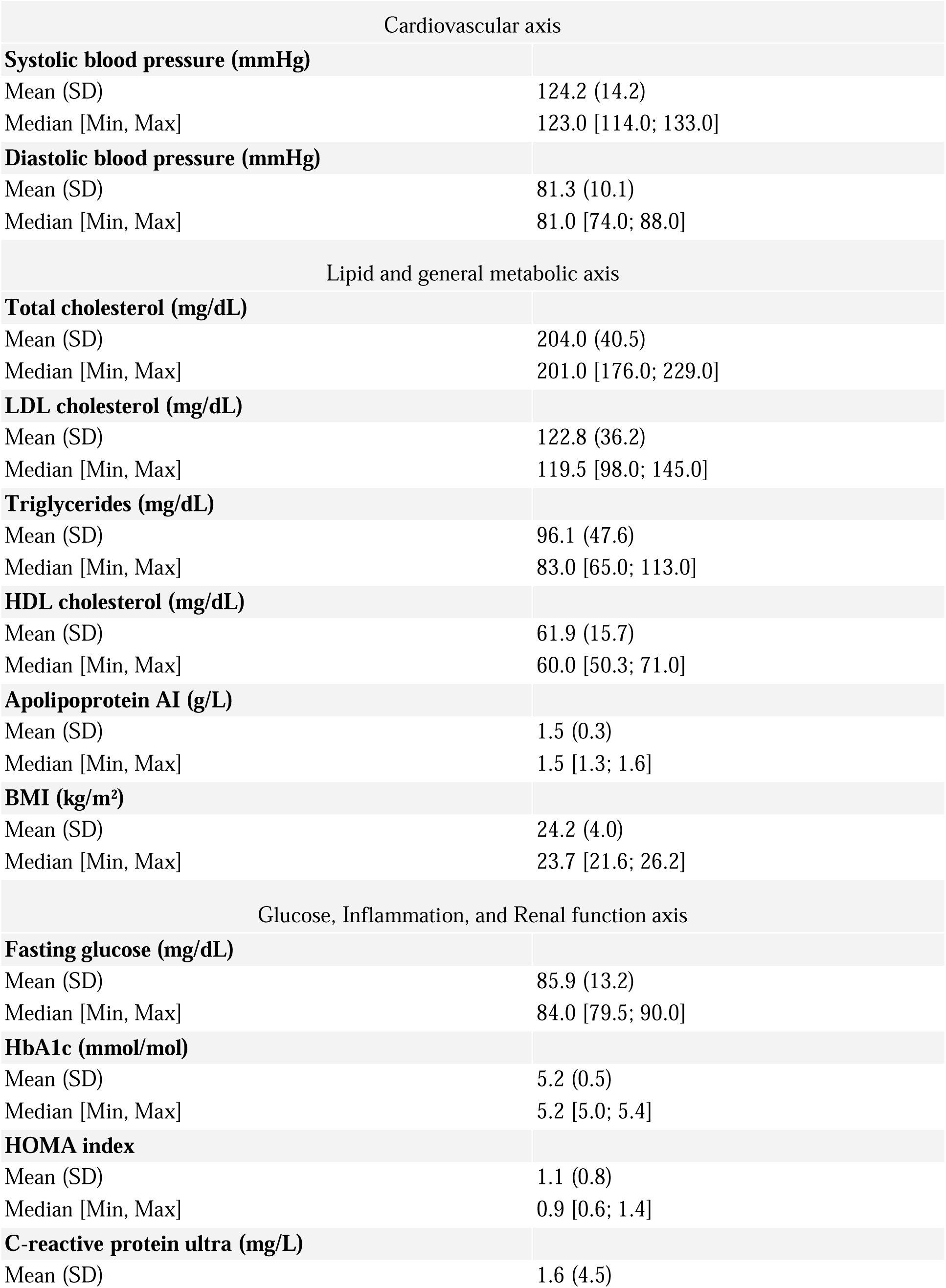
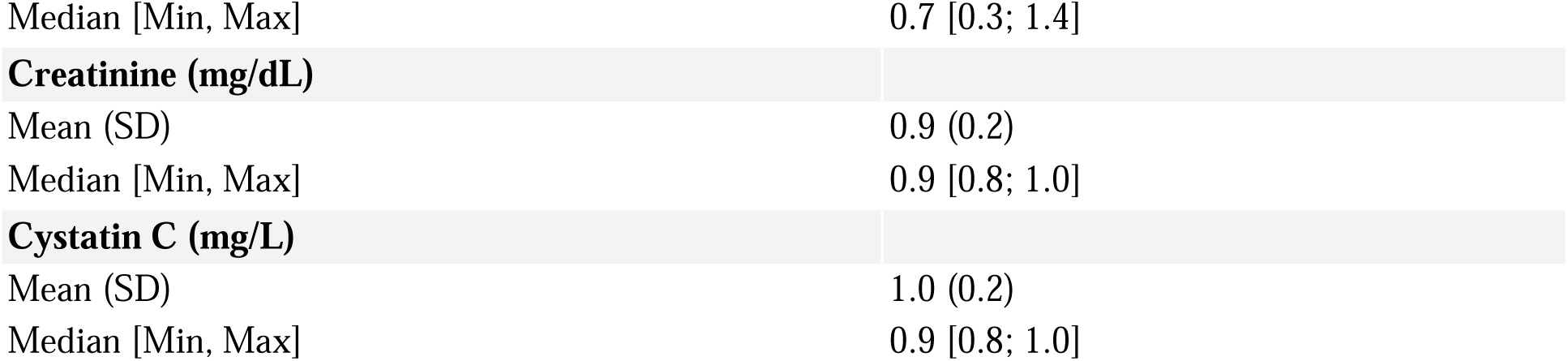
Baseline characteristics of the full study population (N = 1,383) (A) Sociodemographic and anthropometric characteristics; (B) Biomarker distribution across physiological domains used for allostatic load computation

**A. Sociodemographic and anthropometric characteristics**
|  | **Total**<br>**(N=1,383)** |
| --- | --- |
| **Sex** |  |
| Female | 470 (34%) |
| Male | 913 (66%) |
| **Age (years)** |  |
| Mean (SD) | 49 (12.1) |
| Median [Min, Max] | 50 [21; 91] |
| **Education level** |  |
| Primary education (before high school) | 52 (3.7%) |
| Secondary education (high school degree) | 99 (7.1%) |
| Undergraduate degree | 219 (15.7%) |
| Higher education (graduate or above) | 1008 (72.3%) |
| Missing or don't want to answer | 16 (1.2%) |
| **Smoking** |  |
| Yes | 188 (13.5%) |
| No | 1191 (86.2%) |
| n/a | 4 (0.3%) |
| **Body Mass Index (kg/m<sup>2</sup>)** |  |
| Mean (SD) | 24.2 (3.9) |
| Median [Min, Max] | 23.7 [21.6; 26.2] |
| **Body Mass Index status** |  |
| Normal weight or underweight | 878 (63.1%) |
| Overweight | 404 (29.1%) |
| Obesity | 109 (7.8%) |
| Obesity class 1 | 91 (6.5%) |
| Obesity class 2 | 15 (1.1%) |
| Obesity class 3 | 6 (0.4%) |
| **Measured hypertension** |  |
| Yes | 331 (23.9%) |
| No | 1052 (76.1%) |
| B. Biomarker distribution across physiological domains used for allostatic load computation |  |
| Hypothalamic–Pituitary–Adrenal (HPA) axis |  |
| **Evening cortisol (nmol/L)** |  |
| Mean (SD) | 1.4 (2.0) |
| Median [Min, Max] | 0.7 [0.1; 1.8] |
| **Cortisol at awakening (nmol/L)** |  |
| Mean (SD) | 8.2 (4.8) |
| Median [Min, Max] | 7.5 [5.1; 10.4] |
| **Cortisol 30 minutes post-awakening (nmol/L)** |  |
| Mean (SD) | 10.9 (5.1) |
| Median [Min, Max] | 10.3 [7.4; 13.8] |
| **Serum Dehydroepiandrosterone (DHEA) (µg/dL)** |  |
| Mean (SD) | 151.2 (90.3) |
| Median [Min, Max] | 134.0 [84.0; 203.0] |
| Sympathetic Nervous System (SNS) axis |  |
| **Urinary norepinephrine-to-creatinine ratio** |  |
| Mean (SD) | 31.3 (13.6) |
| Median [Min, Max] | 28.6 [22.3; 37.5] |
| **Epinephrine-to-creatinine ratio** |  |
| Mean (SD) | 5.9 (4.2) |
| Median [Min, Max] | 5.0 [3.4; 7.3] |
| **Urinary dopamine (µg/L)** |  |
| Mean (SD) | 167.6 (279.9) |
| Median [Min, Max] | 150.0 [124.0; 185.0] |

| Cardiovascular axis |  |
| --- | --- |
| **Systolic blood pressure (mmHg)** |  |
| Mean (SD) | 124.2 (14.2) |
| Median [Min, Max] | 123.0 [114.0; 133.0] |
| **Diastolic blood pressure (mmHg)** |  |
| Mean (SD) | 81.3 (10.1) |
| Median [Min, Max] | 81.0 [74.0; 88.0] |
| Lipid and general metabolic axis |  |
| **Total cholesterol (mg/dL)** |  |
| Mean (SD) | 204.0 (40.5) |
| Median [Min, Max] | 201.0 [176.0; 229.0] |
| **LDL cholesterol (mg/dL)** |  |
| Mean (SD) | 122.8 (36.2) |
| Median [Min, Max] | 119.5 [98.0; 145.0] |
| **Triglycerides (mg/dL)** |  |
| Mean (SD) | 96.1 (47.6) |
| Median [Min, Max] | 83.0 [65.0; 113.0] |
| **HDL cholesterol (mg/dL)** |  |
| Mean (SD) | 61.9 (15.7) |
| Median [Min, Max] | 60.0 [50.3; 71.0] |
| **Apolipoprotein AI (g/L)** |  |
| Mean (SD) | 1.5 (0.3) |
| Median [Min, Max] | 1.5 [1.3; 1.6] |
| **BMI (kg/m<sup>2</sup>)** |  |
| Mean (SD) | 24.2 (4.0) |
| Median [Min, Max] | 23.7 [21.6; 26.2] |
| Glucose, Inflammation, and Renal function axis |  |
| **Fasting glucose (mg/dL)** |  |
| Mean (SD) | 85.9 (13.2) |
| Median [Min, Max] | 84.0 [79.5; 90.0] |
| **HbA1c (mmol/mol)** |  |
| Mean (SD) | 5.2 (0.5) |
| Median [Min, Max] | 5.2 [5.0; 5.4] |
| **HOMA index** |  |
| Mean (SD) | 1.1 (0.8) |
| Median [Min, Max] | 0.9 [0.6; 1.4] |
| **C-reactive protein ultra (mg/L)** |  |
| Mean (SD) | 1.6 (4.5) |
| Median [Min, Max] | 0.7 [0.3; 1.4] |
| **Creatinine (mg/dL)** |  |
| Mean (SD) | 0.9 (0.2) |
| Median [Min, Max] | 0.9 [0.8; 1.0] |
| **Cystatin C (mg/L)** |  |
| Mean (SD) | 1.0 (0.2) |
| Median [Min, Max] | 0.9 [0.8; 1.0] |

### Measure of the allostatic load

Allostatic load was computed using a multisystem aggregated approach, consistent with prior works [10, 20, 21]. Five physiological domains were represented by biomarkers available in our cohort. The hypothalamic–pituitary–adrenal (HPA) axis was assessed using cortisol at awakening, cortisol 30 minutes post-awakening, evening cortisol, and serum dehydroepiandrosterone (DHEA). The sympathetic nervous system (SNS) was evaluated through urinary norepinephrine-to-creatinine ratio, epinephrine-to-creatinine ratio, and urinary dopamine. The cardiovascular domain was captured by systolic and diastolic blood pressure. Lipid and general metabolic activity included total cholesterol, LDL cholesterol, triglycerides, HDL cholesterol, apolipoprotein AI, and body mass index. Finally, glucose, inflammation, and renal function were assessed with fasting glucose, HbA1c, HOMA index, C-reactive protein, creatinine, and cystatin C.

Preprocessing : extreme values were winsorized at the 1st and 99th percentiles. Right-skewed variables (e.g., CRP, triglycerides, cortisol measures, catecholamine ratios, HOMA) were log-transformed. Each biomarker was converted to sex-specific z-scores. Protective biomarkers (HDL-cholesterol, apolipoprotein AI, DHEA) were multiplied by −1 so that higher values consistently indicate higher biological risk.

Aggregation: For each domain, we computed the mean of available biomarker z-scores. The overall AL was then defined as the z-score of the mean across the five domain scores. Higher AL indicates greater multisystem physiological burden.

### Perceived stress questionnaire and factor structure

To capture exhaustively perceived stress and related psychological distress, we developed and tested a 34-item self-report questionnaire (**Supplementary Table 1**), with items adapted from established instruments (HAM-A, BAI, STAI, PSS) [22–25] and harmonized to a five-point Likert scale (1 = not at all to 5 = totally). Item wording was oriented so that higher scores reflect worse perceived stress/distress.

We assessed the psychometric structure via exploratory (EFA) and confirmatory (CFA) factor analyses using a random split (two-thirds EFA, one-third CFA). The EFA (maximum likelihood, Oblimin rotation) supported a four-factor oblique solution, with adequate sampling (KMO = 0.96; Bartlett p < 0.001). Items with loadings <0.40 or salient cross-loadings were removed while maintaining ≥3 items per factor. The CFA replicated the four-factor structure with excellent fit (χ²/df = 3.49, CFI = 0.991, TLI = 0.990, RMSEA = 0.0076, SRMR = 0.058). Reliability was high for each factor (Cronbach’s α = 0.85–0.92; McDonald’s ω = 0.89–0.94) and for the global score (α = 0.95; ω = 0.96). Detailed loadings, fit indices, and reliabilities are reported in **Supplementary Table 2** and show very good psychometric qualities.

The final instrument, the Zoī Mental Health Questionnaire (ZMHQ), yields four subscale scores and a global perceived stress score (ZPSS). Although one dimension captures depressive symptoms, the ZPSS is conceptualized as a composite index of perceived stress–related psychological distress, consistent with theoretical models linking chronic stress, affect dysregulation, and allostatic overload [26]. Higher ZPSS indicates higher perceived stress/distress. Global and subscale scores were standardized (z) for analyses.

Emotional expressivity. As a brief proxy of emotional awareness/expressivity, participants answered a single item: “Do you find it easy to express your feelings?”. Response options were Yes or No; missing or skipped responses were coded NA. In order to partially observe the potential impact of personality tendencies that could moderate or explain relationships between AL and perceived stress, we briefly ask participants to report their tendencies to express their emotions: “Do you find it easy to express your feelings?”. As a matter of fact, expression of emotion appears as a great factor of stress feeling /experiences [27].

### Definition of stress subgroups

To examine joint subjective–biological patterns, we classified participants into four a priori subgroups based on the distribution of standardized allostatic load (AL) and perceived stress (ZPSS). Following our conceptual framework, these four profiles correspond to distinct configurations of biological and psychological stress:

- Homeostatic profile (ZPSS↓ & AL↓): ZPSS < median and AL < 75th percentile
- Prodromal profile (ZPSS↑ & AL↓): ZPSS ≥ median and AL < 75th percentile
- Latent profile (ZPSS↓ & AL↑): ZPSS < median and AL ≥ 75th percentile
- Decompensated profile (ZPSS↑ & AL↑): ZPSS ≥ median and AL ≥ 75th percentile

This operational rule mirrors prior dissociation work and emphasizes high multisystem burden (upper quartile of AL) versus typical physiological levels, while allowing clear differentiation between psychological and biological stress components.

### Statistical analysis

All analyses were performed in R (RStudio 2025.05.1+513). Continuous variables are summarized as mean ± SD or median [IQR]; categorical variables as counts (percentages). Two-sided tests were used throughout with α = 0.05. For between-group comparisons: Welch’s t-test or Wilcoxon rank-sum test (continuous) and Chi-square test (categorical). For >2 groups: Welch’s ANOVA or Kruskal–Wallis, followed when the omnibus p < 0.05 by pairwise post-hoc tests with Benjamini–Hochberg FDR correction. Effect sizes were reported as Hedges’ g and Cliff’s delta for continuous variables and Cramer’s V for categorical variables. To assess the coupling between multisystem biology and perceived stress, we estimated partial correlations between AL and ZPSS (global and subscales) using linear-model residualization adjusted for age, sex, and BMI, with robustness checks via rank-based (Spearman) and 5% trimmed analyses. Analyses were confirmatory with respect to the pre-specified hypothesis of dissociation between self-reported and biological stress documented in prior literature. For emotional expressivity, we cross-tabulated Yes/No across the four stress profiles and performed a Pearson Chi-square test on complete cases (NA excluded). We report the omnibus statistic (χ², df, p-value) and standardized Pearson residuals for each cell to identify which groups/layers drive the association (positive residuals indicate higher-than-expected counts, negative residuals lower-than-expected).

Quality control. Units and sample handling followed standard clinical procedures (e.g., catecholamines normalized to urinary creatinine; hs-CRP assays). Sensitivity analyses varying the AL domain-completion threshold (≥40–60%) and the AL high-burden cut-point (Q70–Q80) yielded similar subgroup prevalences and inferences (available upon request).

## Results

### Population characteristics

We analyzed 1,383 adult participants from the Zoī cohort. The mean age was 49 ± 12.1 years, 33.9% were women (470/1,383), and the mean BMI was 24.2 ± 4.0 kg/m². This full cohort was used for the development and psychometric validation of the Zoī Mental Health Questionnaire (ZMHQ) and the derivation of the global perceived stress score (ZPSS). Participants underwent an extensive health evaluation covering cardiovascular, neuroendocrine, metabolic, renal, and inflammatory systems. Descriptive statistics are provided in **Table 1**.

### Correlation between AL and ZPSS

Among the 1,383 participants, 1,361 had complete data for both perceived stress (ZPSS) and multisystem allostatic load (AL), and were included in the correlation analyses.To test a replication hypothesis of prior reports suggesting weak coupling between biological and perceived stress, we examined the association between multisystem allostatic load and perceived stress (ZPSS). The partial correlation between standardized AL and the 4-factor ZPSS total, adjusted for age, sex, and BMI, was weak and negative (r ≈ −0.05; n ≈ 1,300), indicating minimal shared variance. A robust Huber regression yielded a similar slope, suggesting the result was not driven by outliers (**Supplementary Figure S1**).

Secondary analyses of AL domains and individual biomarkers with ZPSS dimensions identified some nominal associations, but none remained significant after multiple-testing correction (Benjamini–Hochberg). Overall, perceived stress and allostatic load appeared largely uncoupled in this population (**Supplementary Table 3**).

### Characterization of stress subgroups

Participants were partitioned into four pre-specified stress profiles using the joint distribution of multisystem allostatic load (AL) and perceived stress (ZPSS) in the complete-case sample (n = 1,362) drawn from the full cohort (N = 1,383). We defined a homeostatic profile (ZPSS↓ & AL↓; n = 493; 36.2%), a prodromal profile (ZPSS↑ & AL↓; n = 527; 38.7%), a latent profile (ZPSS↓ & AL↑; n = 188; 13.8%), and a decompensated profile ZPSS↑ & AL↑; n = 154; 11.3%) (**Figure 1**).

**Figure 1.**
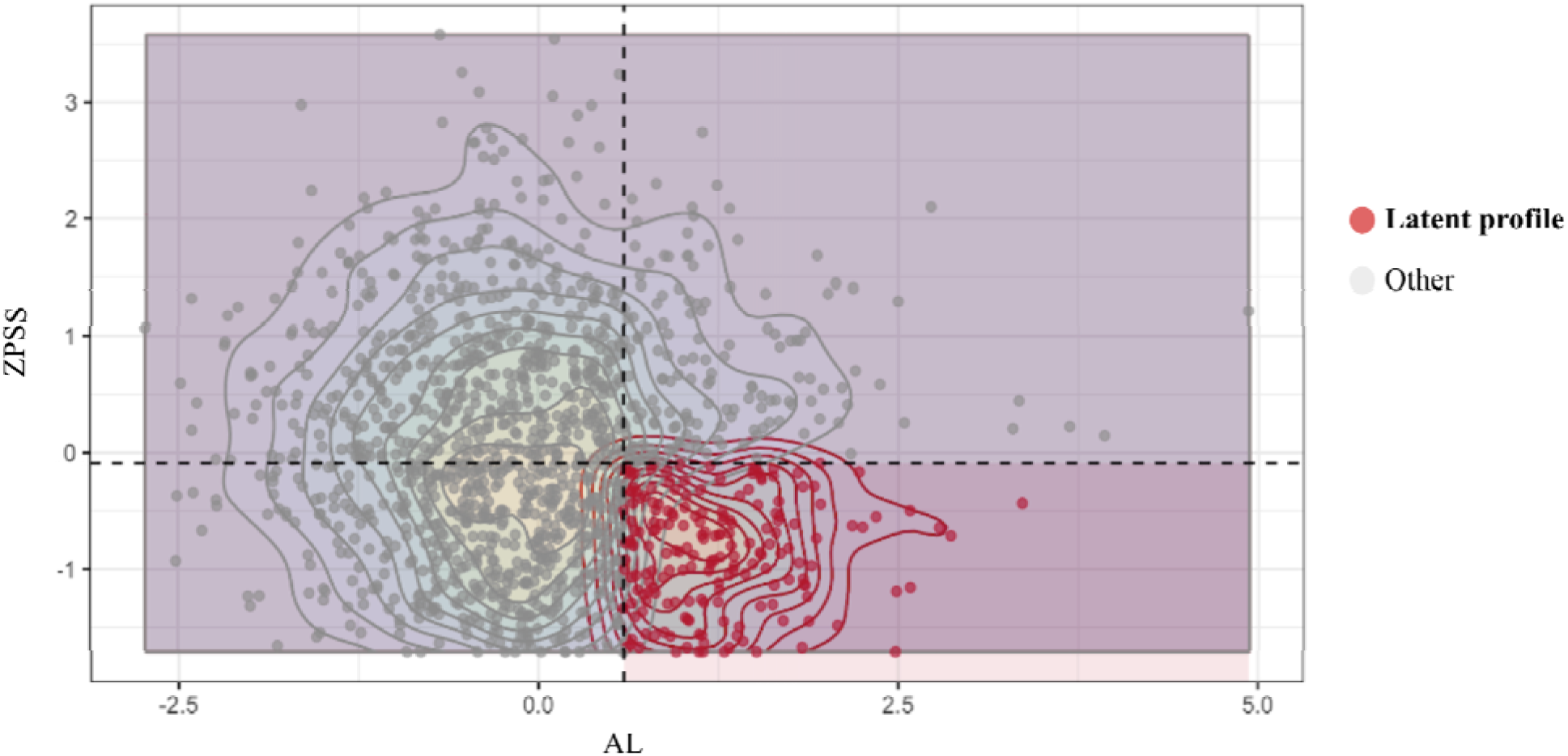
Distribution of participants across stress subgroups. Scatterplot of perceived stress (ZPSS) versus multisystem allostatic load (AL), highlighting the four subgroups, with a focus on the asymptomatic quadrant (AL ≥ Q75 and ZPSS ≤ median).

Omnibus tests confirmed significant group differences across all five domains (Cardiovascular, Glucose–Inflammation–Renal, HPA, Lipid/Metabolic, and SNS; all q < 0.001). Domain-specific scores are shown in **Figure 2** and **Supplementary Table 4**.

**Figure 2.**
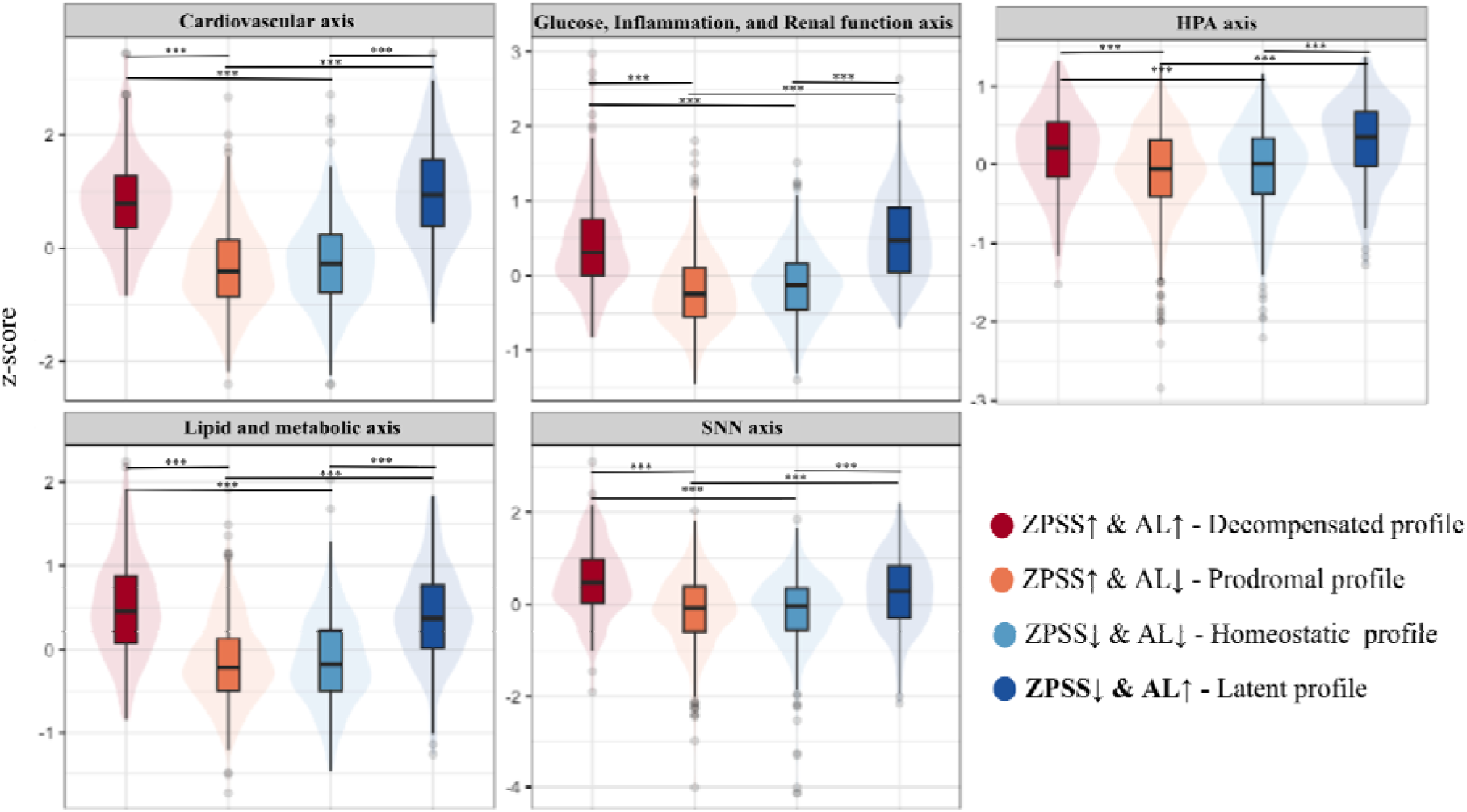
Domain-specific AL z-scores across subgroups. Violin plots showing domain-level scores (Cardiovascular, Glucose Inflammation Renal, HPA, Lipide & Metabolic, SNS) across the four stress subgroups.Only comparisons with extreme significance (q < 0.001) are displayed and marked with ***.

#### Homeostatic profile (ZPSS↓ & AL↓)

This was the largest subgroup, characterized by low scores across all domains. No significant difference were observed compared with the prodromal profile (q > 0.1 across domains), suggesting comparable biological homeostasis despite different subjective experiences of stress.

#### Prodromal profile (ZPSS↑ & AL↓)

These individuals displayed higher perceived distress but physiological scores similar to the homeostatic group, and markedly lower than latent or decompensated participants (q < 1e-25 for Cardiovascular, Glucose–Inflammation–Renal, and Lipid/Metabolic domains). This dissociation reinforces that psychological stress can occur without sustained biological dysregulation.

#### Latent profile (ZPSS↓ & AL↑)

Compared with the homeostatic profile, these participants showed significantly higher scores across all domains (all q < 1e-12). When compared with the decompensated profile, differences were more limited, with divergence restricted to the HPA (q = 0.069) and SNS (q = 0.007) domains.

#### Decompensated profile (ZPSS↑ & AL↑)

Participants with both high perceived stress and high biological load exhibited the most consistent pattern of multisystem dysregulation, with elevated scores across all domains (all q < 0.001).

These results reveal a sizeable subgroup (≈14%) with objectively high multisystem allostatic load despite low perceived stress (latent profile). Nearly half of the cohort exhibited either elevated perceived or biological stress, underscoring the high prevalence of stress-related dysregulation even in an otherwise healthy preventive-medicine population.

### Additional self-report measures

We further explored whether emotional expressivity differed across stress profiles, using the item “Do you find it easy to express your feelings?”. A significant association was found (χ² = 30.2, df = 3, p < 0.001; **Supplementary Table 5**). The latent profile (ZPSS↓ & AL↑) reported greater ease in expressing their emotions (72%) compared with all other groups, whereas both high-perceived-stress profiles, prodromal (ZPSS↑ & AL↓) and decompensated (ZPSS↑ & AL↑), were more likely to respond “No” (44% and 48%, respectively). The homeostatic profile (ZPSS↓ & AL↓) showed intermediate values (67% “Yes”). Standardized residuals indicated that latent participants reported “Yes” responses more often than expected (residual +1.9), while high-perceived-stress profiles showed a relative excess of “No” responses (residuals ≈ +2.1).

## Discussion

This study identifies and confirms a reproducible typology of stress adaptation based on the joint distribution of perceived and biological stress. Among the 1,383 participants from the Zoī preventive cohort, nearly half exhibited either elevated perceived or physiological stress, highlighting the widespread presence of stress-related dysregulation even in an ostensibly healthy population. Within this framework, four distinct profiles emerged, homeostatic, prodromal, latent, and decompensated, representing complementary modes of stress adaptation across subjective and biological dimensions. Our findings resonate with the unified framework proposed by Epel et al. [13], which argues that stress cannot be reduced to a single dimension, biological, psychological, or social, but should be understood through their dynamic interplay. By operationalizing both perceived and physiological stress, our four-quadrant typology provides an empirical instantiation of this integrated model, revealing how discordant responses (prodromal and latent profiles) emerge within a single population.

### Homeostatic profile (ZPSS↓ & AL↓)

The homeostatic quadrant was the largest group, combining low perceived and biological stress. Participants showed the most favorable cardiometabolic profiles and balanced neuroendocrine regulation, reflecting an effective allostatic system. They also reported high emotional expressivity (67% answered “Yes” to “Do you find it easy to express your feelings?”), consistent with adaptive emotion regulation and greater awareness of internal states. This aligns with models describing emotional expressivity and positive affect as protective factors that support autonomic flexibility and parasympathetic recovery. Such individuals maintain low allostatic burden despite environmental challenges, possibly supported by healthy behaviors, social support, and emotional awareness [9, 28].

### Prodromal profile (ZPSS↑ & AL↓)

The prodromal group reported high subjective stress but showed no measurable biological overload. This pattern may correspond to overappraisal of threat or cognitive–emotional hypersensitivity, where individuals experience strong emotional distress without corresponding physiological activation. Participants in this quadrant also tended to report greater difficulty expressing emotions (43.6% “No”), suggesting that emotional suppression or rumination might amplify stress perception while limiting physiological discharge. This echoes prior work linking maladaptive emotion regulation to exaggerated perceived stress and anxiety without corresponding endocrine activation [29]. Clinically, this group could benefit from psychological or behavioral interventions targeting cognitive reframing and emotion regulation before biological dysregulation sets in [30, 31].

### Decompensated profile (ZPSS↑ & AL↑)

The decompensated quadrant combines both high perceived and biological stress, the classical presentation of chronic stress. These participants exhibited the highest multisystem AL, encompassing dysregulation across HPA, cardiovascular, metabolic, and inflammatory domains. Nearly half (48%) reported difficulty expressing emotions, consistent with patterns of emotional inhibition described in burnout and chronic stress conditions [32]. This group likely represents individuals in the advanced stage of stress maladaptation, characterized by sustained HPA axis activation, autonomic imbalance, and pro-inflammatory signaling [33]. Biological features such as flattened cortisol rhythm, reduced heart-rate variability, and elevated cytokines have been reported as key markers of this transition from adaptation to decompensation [34].

### Latent/asymptomatic profile (ZPSS↓ & AL↑)

The latent/asymptomatic quadrant represents the reverse dissociation: elevated biological stress with low perceived stress. Approximately 14% of participants belonged to this group. Notably, they also exhibited the highest emotional expressivity (72% “Yes”, vs. χ² = 30.20, p < 0.001 across groups), suggesting that these individuals find it easy to express their emotions yet still accumulate physiological strain.

Several mechanisms may explain this paradoxical dissociation. Emotional expressivity may buffer subjective stress by facilitating cognitive reappraisal or social regulation of emotion, thereby lowering perceived distress while leaving underlying physiological activation largely unchanged. This interpretation aligns with expressive buffering models, in which verbal or behavioral expression reduces the felt experience of stress without necessarily attenuating neuroendocrine or autonomic responses.

In addition, repeated stress exposure may produce partial psychological habituation, whereby subjective stress responses diminish more quickly than biological ones. Experimental meta-analyses of repeated psychosocial stress show robust habituation of HPA responses but inconsistent or incomplete habituation across autonomic and inflammatory pathways [35], a pattern that closely mirrors the dissociation observed here.

A further possibility is that these individuals carry physiological imprints of past or cumulative stress exposures, including traumatic events. Extensive research in trauma and chronic stress demonstrates that HPA-axis alterations, autonomic imbalance, inflammatory activation, and epigenetic modifications can persist for years after subjective distress has resolved [36–39]. Under this interpretation, high AL in the latent profile may reflect residual or “scar-like” biological signatures of earlier stress, even in the absence of current perceived stress.

Finally, this subgroup may represent an early physiological pre-burnout state [40, 41]. Individuals may maintain emotional and behavioral functioning at the cost of silent biological wear, consistent with reports of subclinical inflammation, altered cortisol rhythm, and reduced heart-rate variability as early markers of physiological overload [33].

The latent pattern also contrasts with the alexithymic phenotype (low expressivity, high physiological reactivity [42, 43]). Together, these findings suggest that dissociation between emotional appraisal and physiological load can arise through different pathways one marked by emotional unawareness, the other by expressive buffering or over-adaptation. Although limited by the single-item measure of emotional expressivity, these results highlight how emotional processing shapes perceived stress without necessarily protecting against multisystem biological burden. Future work using validated multidimensional measures (TAS-20, ERQ, MAIA) will be needed to disentangle these mechanisms more precisely.

*Integrative perspective* Altogether, these findings depict a continuum of stress adaptation from homeostasis to decompensation, with two intermediate dissociative modes, the prodromal and latent profiles, each reflecting a partial failure of mind–body synchronization. Mechanistically, both dissociations likely involve altered interoception, central regulation, and limbic–autonomic coupling [16]. Experimental work on repeated psychosocial stress also suggests that individual differences in the habituation of biological systems may underlie such dissociations. A recent meta-analysis of repeated TSST protocols reported robust habituation of HPA responses but more variable and often incomplete habituation of autonomic and inflammatory markers [35]. Our high-AL profiles, particularly the latent subgroup, may therefore reflect insufficient biological habituation despite a blunted subjective experience of stress.

The mismatch between subjective and physiological stress is further supported by experimental studies demonstrating that perceived stress and biological activation can become temporally desynchronized through long-term allostatic adaptation [44]. Similar dissociations have been documented in post-traumatic stress research, where biological signatures of stress persist long after subjective distress has diminished. PTSD studies show durable alterations in HPA-axis regulation, autonomic balance, inflammatory signaling, and epigenetic patterns despite partial or full psychological remission [37–39]. These findings support the notion that elevated AL may reflect past or cumulative stress exposures even when current perceived stress is low. In the long term, such “silent” stress phenotypes may accelerate biological aging, as chronic stress contributes to telomere shortening, mitochondrial dysfunction, and systemic inflammation [45].

*Implications for preventive medicine* Detecting stress-related dysregulation in a population as health-conscious as the Zoī cohort [19] underscores that biological stress imbalance can emerge independently of lifestyle risk factors. Our typology further shows that a substantial fraction of individuals fall into prodromal (high perceived, low biological stress) or latent (low perceived, high biological stress) profiles, which would remain largely undetected by routine clinical assessment. These findings strengthen the case for integrating biological stress indices (e.g., AL, HRV, cytokines) into preventive health programs, as reliance on self-reported stress alone would miss a substantial fraction of at-risk individuals [8, 34, 35].

Furthermore, biosensors and ecological momentary assessments now enable real-time monitoring of physiological stress markers such as HRV, electrodermal activity, or cortisol, supporting scalable detection of subclinical stress in daily life [4, 31, 36]. These directions align with the French National Mental Health and Psychiatry Plan 2025–2030, HAS, 2025, which emphasizes early identification of physiological dysregulation as a foundation for predictive, personalized prevention.

Within the Zoī framework, combining AL with oxidative stress markers, inflammatory cytokines (IL-6, TNF-α, hs-CRP), and imaging-derived phenotypes could refine stratification and improve prediction of outcomes such as metabolic syndrome, depression, or cardiovascular disease [37, 38]. Such multimodal, data-driven models may ultimately enable clinicians to identify individuals in pre-symptomatic but biologically stressed states, particularly those in the latent profile, opening the way to early, targeted, and emotion-informed interventions.

## Limitations

First, the cross-sectional design precludes causal inference. We cannot determine whether biological dysregulation (high AL) precedes changes in perceived stress or represents a compensatory state. Longitudinal follow-up is needed to clarify whether the prodromal and latent profiles correspond to transient adaptations, stable traits, or early stages of pre-burnout progression [15, 16]. In addition, allostatic load may partly reflect past or cumulative stress exposures, including traumatic events, whose physiological imprints can persist long after subjective distress has diminished. Conversely, high perceived stress in the absence of elevated AL may reflect more recent or acute stress that has not yet left a measurable biological trace [51]. This temporal decoupling between perceived and biological stress should be explicitly considered when interpreting dissociative profiles.

Second, the perceived stress instrument (ZPSS/ZMHQ), although internally consistent and psychometrically validated within this cohort, remains an internal tool. External validation against standardized scales such as the Perceived Stress Scale (PSS), Depression Anxiety Stress Scales (DASS), or Toronto Alexithymia Scale (TAS-20) is necessary to confirm its generalizability. In particular, our single-item measure of emotional expressivity provides only a limited proxy for emotional awareness and should be interpreted cautiously. While the significant χ² association (p < 0.001) between stress quadrants and emotional expressivity is compelling, it remains exploratory and requires replication using multi-item and multidimensional instruments (e.g., Emotional Expressivity Scale, ERQ, or MAIA) [45–47]. Moreover, perceived stress and emotional expressivity are influenced by sociocultural norms of emotional reporting (e.g. gender, education, professional role, cultural background), which may systematically bias ZPSS scores independently of biological load. Future work should therefore consider stratified or subgroup analyses to better account for these normative differences.

Third, the operationalization of allostatic load, including biomarker selection, z-score standardization, and domain averaging, represents one of several legitimate modeling strategies. Alternative formulations (e.g., weighted indices, percentile thresholds, or unsupervised clustering) might yield slightly different subgroup boundaries [26, 40]. Similarly, the threshold-based definition of stress subgroups (AL ≥ Q75 and ZPSS ≤ median) is data-driven and should be confirmed in independent samples with prospective outcomes.

Fourth, unmeasured confounders such as medication use (antihypertensives, antidepressants, corticosteroids), menopausal status, circadian sampling time, or recent acute stressors could affect AL or cortisol measures. Sleep disturbances and chronic inflammation, both closely linked to stress physiology, were not systematically assessed.

Finally, while the Zoī cohort provides extensive biological and behavioral coverage, certain relevant systems, such as oxidative stress markers, cytokine panels (IL-6, TNF-α, hs-CRP), and autonomic measures (e.g., HRV), were not yet integrated. Including these parameters will be essential to refine multidimensional modeling of stress biology and identify discriminant markers of homeostatic versus vulnerable profiles [43, 48, 49].

## Perspectives

Future research should pursue longitudinal and mechanistic studies to establish whether the four stress profiles, homeostatic, prodromal, latent, and decompensated, predict differential health trajectories. Tracking individuals over time, with at least three evaluation time points, will clarify whether latent participants evolve toward overt stress or burnout states, whether prodromal participants develop biological dysregulation, and whether emotional expressivity acts as a protective or compensatory mechanism. Rather than fixed categories, these profiles likely represent dynamic positions along a continuum of stress adaptation, with individuals transitioning between states depending on life events, recovery capacity, and cumulative exposures.

A priority will be to develop multimodal discriminant models integrating allostatic load with complementary biomarkers, including oxidative stress indices, inflammatory cytokines, metabolic markers, and imaging-derived phenotypes, to better differentiate stress phenotypes and predict clinical transitions. Machine learning or latent class modeling could help derive reproducible, data-driven typologies beyond threshold-based definitions.

In parallel, psychological and interoceptive dimensions should be more comprehensively captured. Future Zoī investigations could incorporate validated measures of alexithymia (TAS-20), emotion regulation strategies (ERQ), interoceptive awareness (MAIA), perceived control, and coping styles (e.g., cognitive reappraisal, disengagement, social support seeking), together with stress-reactivity tasks and diurnal cortisol sampling. These tools will be critical to test the proposed “expressive dissociation” hypothesis, whereby high emotional expressivity may reduce perceived stress while leaving physiological stress unresolved.

Finally, integrating wearable biosensors and ecological momentary assessments (e.g., heart-rate variability, skin conductance, sleep, mood tracking) will allow real-time monitoring of physiological stress and recovery in naturalistic contexts. Such digital phenotyping, combined with AL and self-report indices, may enable scalable early detection of subclinical stress, particularly in prodromal and latent profiles, and guide personalized, preventive, and emotion-informed interventions.

## Conclusion

By combining perceived and biological stress assessments, this study delineates a four-quadrant typology of stress adaptation, encompassing homeostatic, prodromal, latent, and decompensated profiles. Notably, a non-negligible latent phenotype emerged, characterized by high allostatic load in the absence of reported distress. Overall, a substantial proportion of this preventive cohort exhibited either psychological or physiological stress, underscoring the pervasiveness of stress-related dysregulation even among otherwise healthy individuals.

Emotional expressivity appeared to modulate perceived stress without fully protecting against physiological strain, suggesting that emotional awareness may buffer subjective experience more effectively than underlying biology. Recognizing, modeling, and longitudinally tracking these profiles, particularly the prodromal and latent subgroups, may help reorient preventive and precision medicine toward earlier, multimodal, and emotion-informed interventions before chronic disease onset.

## Data Availability

All data produced in the present study are available upon reasonable request to the authors

## Acknowledgements

The authors express their thanks to Élodie Vorkaufer.

## Consent to participate

The study involved only retrospective, pseudonymized data. All participants were informed generally and individually prior to inclusion, with the possibility to object, in accordance with national regulations.

## Consent to publish

No individual-identifiable data or images are included in this manuscript.

## Ethics approval

The study was approved by the Ethics Committee of *Comité d’Éthique pour la Recherche Biomédicale de Rabat* (CERB 159-25).

